# Early detection of malignant and pre-malignant peripheral nerve tumors using cell-free DNA fragmentomics

**DOI:** 10.1101/2024.01.18.24301053

**Authors:** R. Taylor Sundby, Jeffrey J. Szymanski, Alexander Pan, Paul A. Jones, Sana Z. Mahmood, Olivia H. Reid, Divya Srihari, Amy E Armstrong, Stacey Chamberlain, Sanita Burgic, Kara Weekley, Béga Murray, Sneh Patel, Faridi Qaium, Andrea N. Lucas, Margaret Fagan, Anne Dufek, Christian F. Meyer, Natalie B. Collins, Christine A. Pratilas, Eva Dombi, Andrea M. Gross, AeRang Kim, John S.A. Chrisinger, Carina A. Dehner, Brigitte C. Widemann, Angela C. Hirbe, Aadel A. Chaudhuri, Jack F. Shern

## Abstract

Early detection of neurofibromatosis type 1 (NF1) associated peripheral nerve sheath tumors (PNST) informs clinical decision-making, potentially averting deadly outcomes. Here, we describe a cell-free DNA (cfDNA) fragmentomic approach which distinguishes non-malignant, pre-malignant and malignant forms of NF1 PNST. Using plasma samples from a novel cohort of 101 NF1 patients and 21 healthy controls, we validated that our previous cfDNA copy number alteration (CNA)-based approach identifies malignant peripheral nerve sheath tumor (MPNST) but cannot distinguish among benign and premalignant states. We therefore investigated the ability of fragment-based cfDNA features to differentiate NF1-associated tumors including binned genome-wide fragment length ratios, end motif analysis, and non-negative matrix factorization deconvolution of fragment lengths. Fragmentomic methods were able to differentiate pre-malignant states including atypical neurofibromas (AN). Fragmentomics also adjudicated AN cases suspicious for MPNST, correctly diagnosing samples noninvasively, which could have informed clinical management. Overall, this study pioneers the early detection of malignant and premalignant peripheral nerve sheath tumors in NF1 patients using plasma cfDNA fragmentomics. In addition to screening applications, this novel approach distinguishes atypical neurofibromas from benign plexiform neurofibromas and malignant peripheral nerve sheath tumors, enabling more precise clinical diagnosis and management.

## Introduction

Neurofibromatosis type 1 (NF1) is the most common heritable cancer predisposition syndrome and is characterized by a spectrum of benign, pre-malignant, and malignant nerve sheath tumors. Approximately 50% of patients with NF1 develop benign plexiform neurofibromas (PN)^1^, typically present in infancy or early childhood^2^, with a subset of PN evolving into pre-malignant atypical neurofibromas (AN)^3–5^ and, ultimately, 8-15% of patients with NF1 developing cancerous malignant peripheral nerve sheath tumors (MPNST) during their lifetime^6–8^. MPNST account for the majority of NF1-associated mortality^6,7^ with a 5-year overall survival rate of only 20%^9^. Genomic and histopathologic evidence suggests a model in which PN evolve to MPNST through an intermediary AN disease state^3^ with clinical evidence of AN lesions transforming directly into MPNST^4^.

Diagnostically, however, differentiating between PN, AN, and MPNST remains clinically challenging as a result of insensitive clinical exams^10,11^, overlapping findings on imaging^12^, and tissue heterogeneity leading to sampling biases on tissue biopsy^13^. Biopsy also carries with it the risk of peripheral nerve injury^14,15^, further complicating the diagnostic workup. This is unfortunate as the therapeutic management of these entities is quite different, given their varying levels of malignant potential. PN are typically observed or treated with mitogen activated protein kinase/ extracellular signal regulated kinase kinase (MEK) inhibitors^16,17^; AN are typically surgically removed with narrow margins^4,15,18,19^; MPNST require more morbid wide margin resections to prevent recurrence^19^, yet are often metastatic at diagnosis^20^.

We previously demonstrated that a liquid biopsy approach utilizing cell-free DNA (cfDNA) copy number alteration (CNA) analysis accurately distinguishes MPNST from benign PN, and that mean cfDNA fragment length is shorter in MPNST than in PN patients or healthy volunteers^21^. In the current study, we significantly extend these findings to demonstrate early cancer and pre-cancer detection from plasma cfDNA by noninvasively distinguishing between premalignant tumor cell states. To achieve specificity for different premalignant states, we quantify several fragment-level cfDNA features including bin-wise analysis of short and long cfDNA fragment ratios, deconvolution of fragment end motif profiles, and deconvolution of fragment length profiles using non-negative matrix factorization (NMF). While each method provided complementary diagnostic data, NMF of fragment length profiles performed best at granularly distinguishing between malignant, pre-malignant and non-malignant states. Sophisticated fragmentomic analysis of cfDNA therefore has the potential to distinguish between non-cancer, pre-cancer, and cancer states and facilitate early cancer and pre-cancer screening for the most deadly malignancy associated with NF1.

## Results

### Copy number alterations in cell-free DNA identify malignant peripheral nerve sheath tumors but do not differentiate pre-malignant states

In this study we sought to determine whether features of cfDNA could distinguish between benign (PN) and pre-malignant (AN) states in the cancer predisposition syndrome NF1. To address this question, we collected samples from a cohort of participants with MPNST (n = 35), treated MPNST with no evidence of disease (NED) (n = 15), MPNST on treatment (n = 10), PN (n = 69), or AN (n = 17), and healthy controls (n = 21) (**Supplemental Tables 1-3**). cfDNA from these participants was prepared and sequenced under standardized conditions to allow for fragment length and end motif analysis (8 PCR cycles, 6x depth, see **Methods**).

Previously, using a separate cohort, we demonstrated that *in silico* size-selected plasma cfDNA CNA can distinguish MPNST from PN or healthy states^21^. Our new participant cohort validates this finding comparing MPNST to PN (AUC = 0.73) and to healthy controls (AUC = 0.80). Tumor fraction also distinguished AN from MPNST (AUC = 0.75) (**Figure 1 a-b**). Additionally, the new cohort validated that healthy controls’ fragment length distributions differ from PN (D = 0.0241, p < 0.001 by two-sample Kolmogorov–Smirnov (KS) test) and MPNST (D = 0.0138, p < 0.001 by two-sample KS test) as well as PN from MPNST (D = 0.0368, p < 0.001 by two-sample KS test). Fragment length distribution was also found to differentiate AN from healthy controls (D =0.0072 p < 0.001 by two-sample KS test), PN (D = 0. 0300, p < 0.001 by two-sample KS test), and MPNST (D = 0. 0.0115, p < 0.001 by two-sample KS test) **(Figure 1c).** However, CNA-derived tumor fraction could not accurately distinguish between non-malignant tumor states (PN vs. healthy, AUC = 0.64; AN vs. healthy, AUC = 0.60; PN vs. AN, AUC = 0.48).

**Figure 1:**
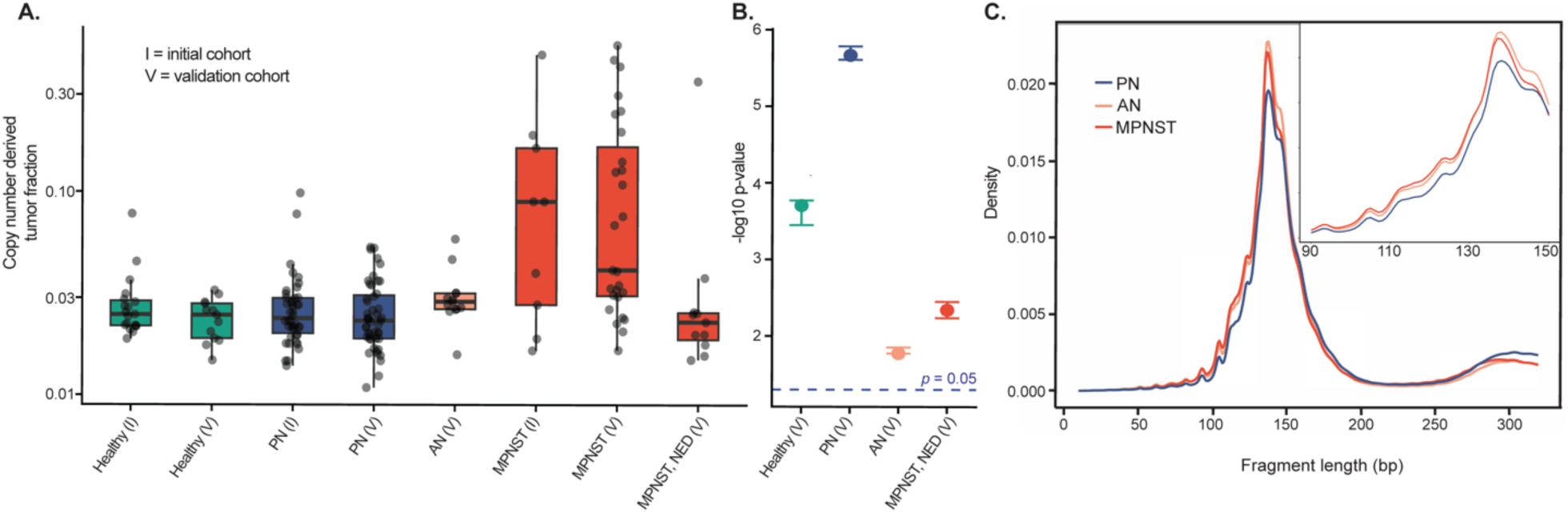
Copy number alterations in cfDNA identify MPNST but cannot resolve premalignant tumor states. (**A**) Highest per-participant size-selected ichorCNA copy-number derived tumor fraction in each clinical state for initial (I) and validation (V) cohorts. (**B**) Significance of validation cohort tumor fraction compared to validation MPNST in leave-one-out Wilcoxon rank-sum tests, expressed as −log10 P values. **(C)** Fragment length densities for cfDNA in NF1 patients with PN, AN, and MPNST peripheral nerve sheath tumors. Colors represent the patient’s clinical diagnosis. AN, atypical neurofibroma; MPNST, malignant peripheral nerve sheath tumor; NED, no evidence of disease; NF1, neurofibromatosis type 1; PN, plexiform neurofibroma. Thus, our validation study confirmed the low diagnostic yield of CNA in cfDNA from participants with low CN-burden PN and AN, but also that these non-malignant tumor states generate distinct cfDNA fragment length profiles (Figure 1c). Therefore, we devised a strategy for comprehensive multi-method characterization of cfDNA fragments in cancer predisposition with the aim of distinguishing all tumor types using only cfDNA (Figure 2a). Building upon our previous findings using fragment size selections to enrich for malignant cfDNA templates, we extended this analysis to examine mapped short-to-long cfDNA fragment ratios, composition of the fragment’s end motifs, and NMF models of fragmentomic feature distributions (Figure 2b).

**Figure 2.**
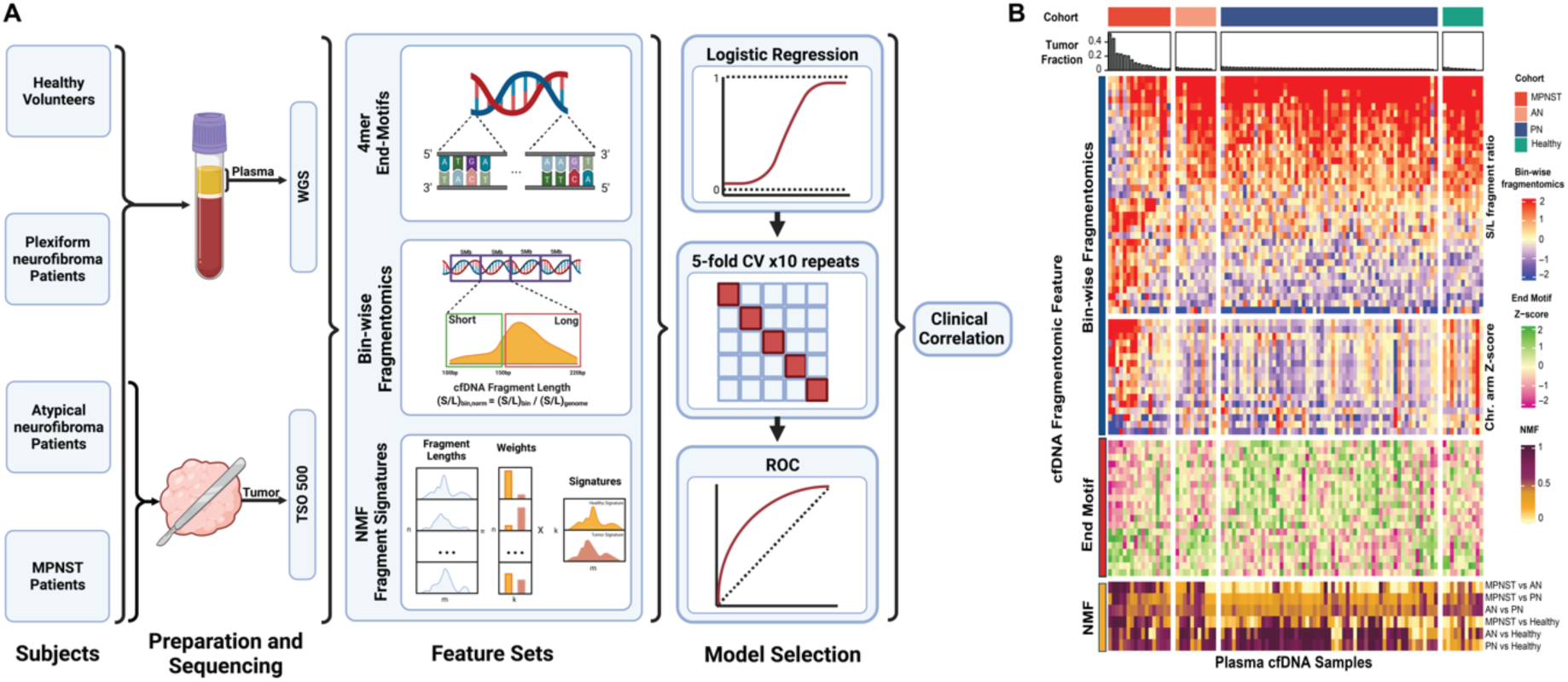
Fragmentomic features differentiate benign, pre-malignant and malignant peripheral nerve sheath tumors. (**A**) Study schema. Participants consisted of patients with imaging- and biopsy-proven MPNST or AN, established patients with PN, and healthy donors. Plasma was collected for fragmentomic analysis. AN and MPNST tissue DNA, when available, was clinically sequenced using the TSO 500 targeted oncology panel. cfDNA was extracted from plasma and underwent whole genome sequencing with fragmentomic profiles assessed by fragment end motifs, bin-wise fragmentomic profiles, and NMF fragment signatures. Models for each feature type were trained on one-versus-one comparisons with resultant features input into a logistic regression with 10 repeats of 5-fold CV. Optimal thresholds were calculated from ROC curve analysis of models’ predicted scores using Youden’s J-statistic. Results were correlated with clinical diagnoses and outcomes. (**B**) Heatmap of fragmentomic features. Samples are grouped by diagnostic cohort (MPNST, AN, PN, healthy) with samples in each cohort ranked from highest to lowest copy number alteration-derived tumor fraction. Rows are grouped by fragmentomic feature type. AN, atypical neurofibroma; bp, base pair; cfDNA, cell-free DNA; CV, cross validation; MPNST, malignant peripheral nerve sheath tumor; NMF, non-negative matrix factorization; PN, plexiform neurofibroma; ROC, receiver operating characteristic; TSO 500, TruSight Oncology 500.

### Genome-wide bin-wise fragmentomics enhance detection of MPNST

Tumor-derived circulating DNA fragments are known to be, on average, shorter than cfDNA from healthy tissues, and bin-wise fragment length ratio has previously been shown to detect a variety of cancers. Consistent with previous reports in other cancer types^22,23^, comparing the ratio of short (<150 bp) to long (>150 bp) cfDNA fragments in 5 Mb bins across the genome (**Methods**), we found substantial aberration in fragmentation profiles of pre-treatment MPNST compared to healthy controls or samples from patients on treatment (**Figure 3a-b**). Globally, fragmentation profiles of PN and AN resembled the healthy state and did not demonstrate the aberrations observed in pre-treatment MPNST. In pairwise comparisons of clinical states, bin-wise fragmentomics differentiated MPNST with high accuracy versus AN, PN, and healthy states (**Figure 3d**). Unlike CNA-derived tumor fraction, fragment length ratio in bin-wise fragmentomics was able to distinguish healthy from PN states (tumor fraction AUC = 0.64, bin-wise fragmentomics AUC = 0.87). Still, performance of bin-wise fragmentomics was low when comparing PN to AN (AUC = 0.59) or AN versus healthy (AUC = 0.45). This method did, however, accurately distinguish AN from MPNST (AUC = 0.75; **Figure 3a**), suggesting this liquid biopsy fragmentomic approach could clinically distinguish between MPNST and its pre- malignant precursor. For example, Subj189 underwent fine-needle biopsy of a tumor for new-onset pain and distinct nodular appearance on MRI of a tumor. Biopsy was consistent with AN, not PN, with atypia and p16 loss on immunohistochemical (IHC) staining suggestive of a *CDKN2A* deletion. Bin-wise fragmentomic one-versus-one (OVO) scores from matched plasma (lib286) were in agreement and consistent with AN (PN-AN 0.74, threshold 0.45; healthy-AN 0.77, threshold 0.63). The subject’s subsequent clinical course, however, was suspicious for comorbid MPNST with 26% growth over a year. Importantly, OVO for AN-MPNST was again consistent with AN, not MPNST (AN-MPNST 0.6, threshold 0.64) and ultimate total resection confirmed the diagnosis of AN without regions of malignant transformation (**Figure 3c**). This case highlights the potential for cfDNA fragmentomics to adjudicate clinically and radiographically equivocal neurofibromas on the malignant vs. pre-malignant spectrum, with important implications for making clinical decisions earlier, more confidently, and more precisely.

**Figure 3.**
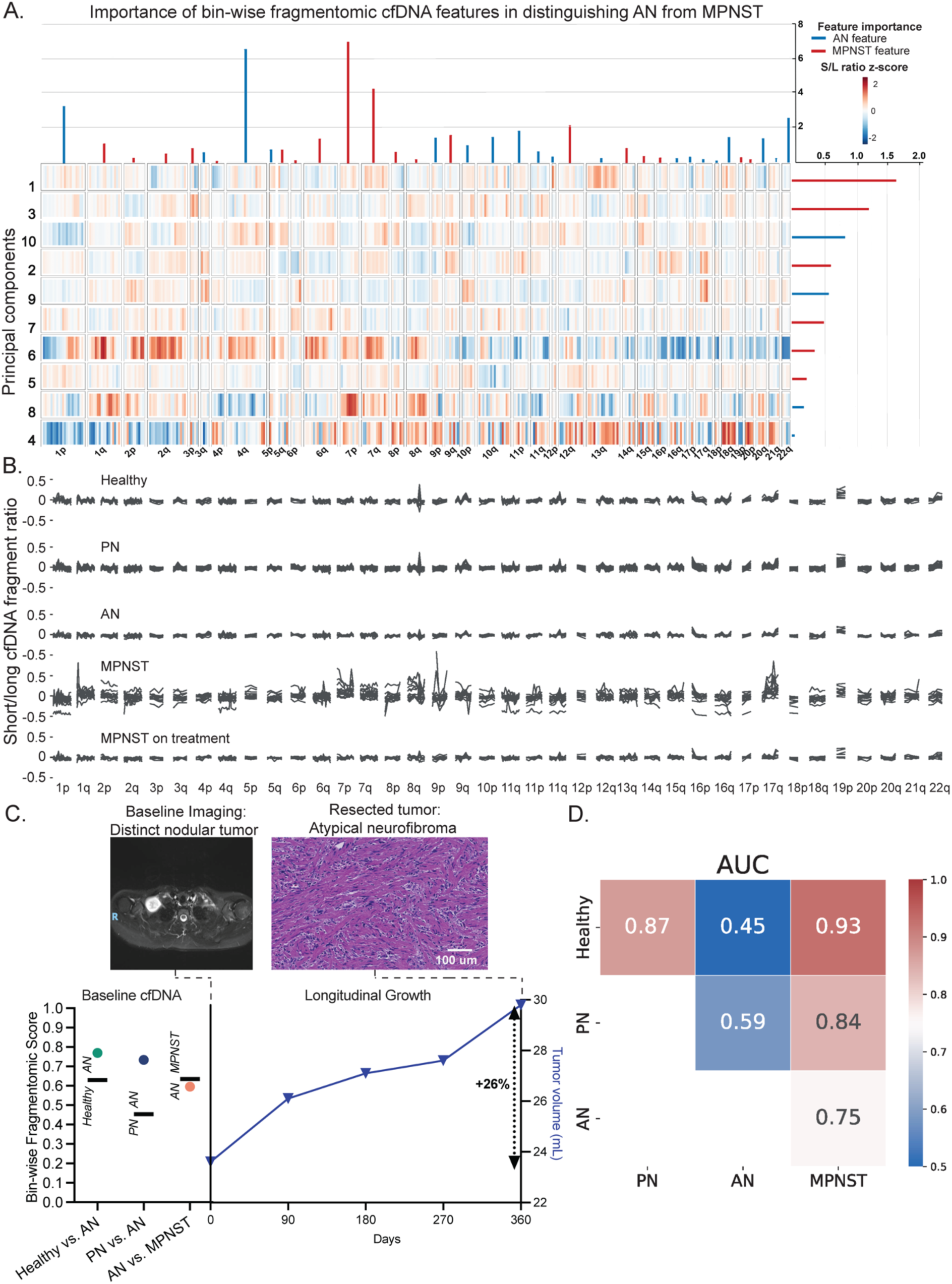
**Bin-wise fragmentomic analysis reveals distinct profiles of healthy controls, PN, and MPNST**. (**A**) Heatmap of principal component eigenvalues of fragmentation profile features differentiating AN from MPNST. The relative importance of the features is represented at the right (bin-wise short/long ratio changes) and top (chromosomal arm changes) of the heatmap. Red feature importance lines indicate features or principal components associated with MPNST, while blue feature importance lines are associated with AN. (**B**) Ratio of short to long fragments in 5 Mb bins across the genome in healthy volunteers and patients with PN, AN, pre-treatment MPNST, and patients receiving treatment for their MPNST. (**C**) Bin-wise fragmentomic scoring and every 3-month surveillance MRI tumor volumes for a patient with AN diagnosed by biopsy, suspicious for malignant transformation given the rate of growth.

Fragmentomic scores for healthy versus AN (green circle), PN versus AN (purple circle), and AN versus MPNST (orange circle) along with discrimination thresholds (horizontal black lines) from a paired blood draw are on the left y-axis. Tumor volumes are on the right y-axis, with 26% growth over 1 year indicated. (**D**) One-versus-one ROC AUCs of logistic regressions with 10 repeats of 5-fold cross validations performed over the bin-wise short/long ratio and chromosomal arm z-score data (**Methods**). Healthy and disease states included within each comparison are shown along x- and y-axes, with AUCs indicated both numerically and by heat level. AN, atypical neurofibroma; AUC, area under the curve; cfDNA, cell-free DNA; MPNST, malignant peripheral nerve sheath tumor; PCA, principal component analysis; PN, plexiform neurofibroma.

### cfDNA fragment end motifs distinguish pre-malignant AN from benign PN and malignant MPNST

cfDNA fragment end motifs have been shown to reflect cfDNA processing and epigenomic profiles with relevance to cancer^24–29^. We therefore hypothesized that, in the setting of NF1 MPNST, certain end motifs are enriched which could facilitate early cancer screening. Indeed, we found that the distribution of end motifs among NF1 associated clinical states was non-random with preferential motifs enriched in one-versus-one comparisons (**Figure 4a**). Still, motif diversity score (MDS), an aggregate measure of motif distribution through normalized Shannon entropy, was not significantly different between clinical states (**Figure 4b-c**). Recently, Zhou et al.^30^ demonstrated that cfDNA fragment cleavage patterns could be quantified by non-negative matrix factorization of 4-mer end motifs into “founder” end motif profiles (F-profiles). We found that indeed, motifs contributed non-randomly to F-profiles (**Supplemental Figure 1**), F-profile contributions to cfDNA samples differed between clinical states (**Figure 4d**), and that specific F- profiles were more accurate than individual motifs in differentiating clinical states. For example, comparing MPNST and AN, F-profile 2 (AUC 0.65) substantially outperformed the most predictive motif, AAAA (AUC 0.59). In differentiating PN versus AN, arguably the most difficult clinical distinction, both specific motif (AUC 0.69) and F-profile (AUC 0.70) performed well compared with the aggregate MDS (AUC 0.48) (**Figure 4e**). Both F-profile 5 and the ACCA motif also out-performed bin-wise fragmentomics (AUC 0.59) when classifying AN vs. PN, showcasing the power of high-resolution fragment end analysis to noninvasively distinguish between pre-malignant neurofibroma states.

**Figure 4.**
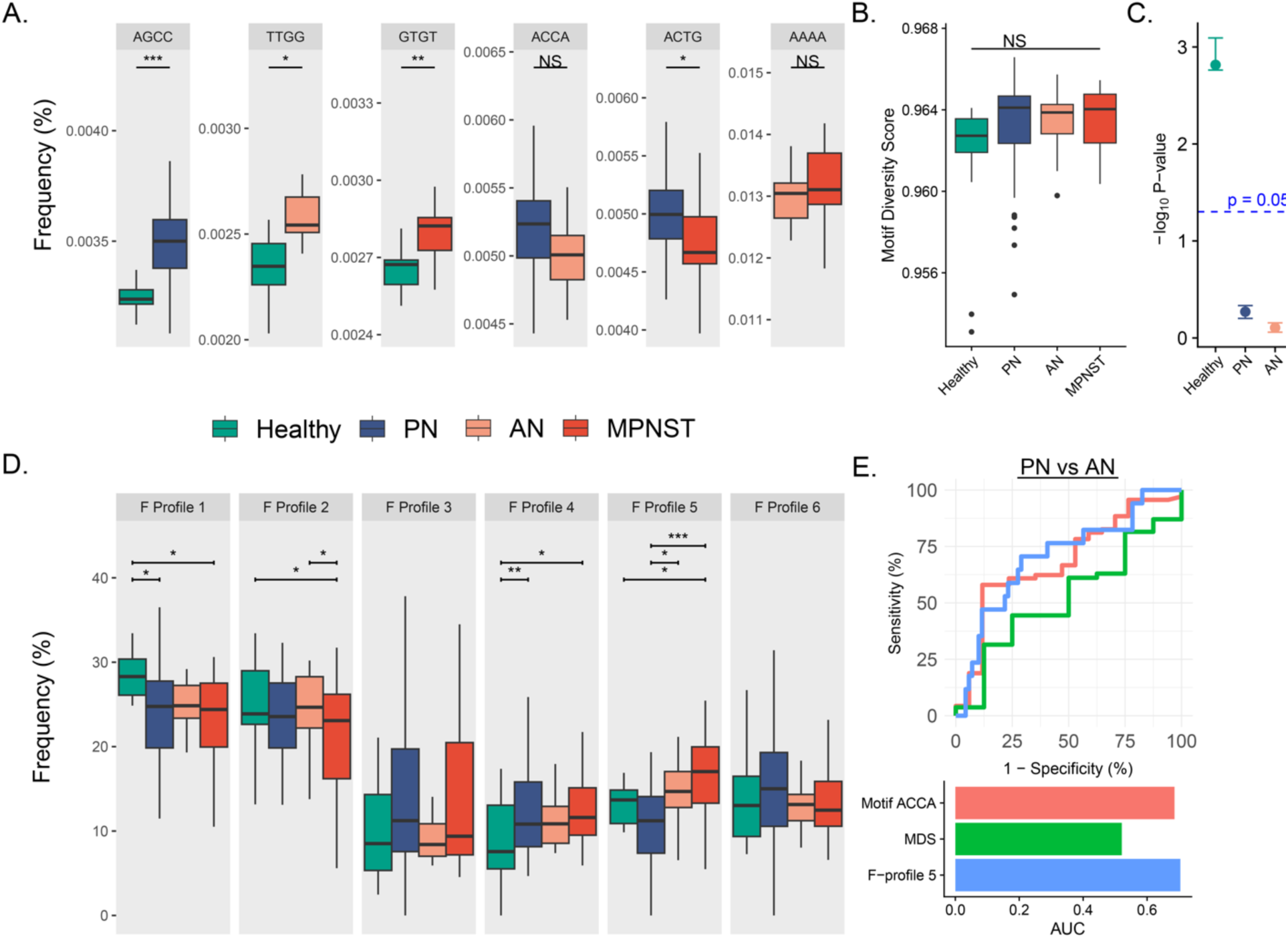
**cfDNA fragment end composition distinguishes pre-malignant from malignant nerve sheath tumors**. (**A**) Specific 4-mer end motifs which best classify between clinical diagnosis pairs in one-versus-one comparisons; (p-values by Benjamini-Hochberg corrected t-test). (**B**) End motif diversity scores (MDS) do not differentiate cohorts; (Kruskal Wallis H-statistic p-value > 0.05), although **(C)** MDS in leave-one-out Wilcoxon rank-sum test against MPNST approach (PN, AN) or surpass (healthy) significance of p < 0.05. **(D)** Percent contribution of NMF-deconvolved end motif profiles (F-profiles) in each plasma cfDNA sample by clinical state (p-values by Tukey post-hoc test after Bonferroni-corrected ANOVA). **(E)** ROC curve comparing AN and plexiform using each fragment end method: best-performing individual end motif (ACCA; AUC = 0.69), motif diversity score (AUC = 0.52), and best distinguishing F-profile (F-profile 5; AUC = 0.70); AN, atypical neurofibroma; cfDNA, cell-free DNA; F-profile, founder end motif profile; MPNST, malignant peripheral nerve sheath tumor; NMF, non-negative matrix factorization; PN, plexiform neurofibroma; ROC, receiver operating characteristic; TF, tumor fraction, * < 0.1; ** < 0.001; *** < 0.0001.

### NMF deconvolves global fragment lengths into disease state specific fragmentomic signatures

We previously published that plasma cfDNA samples from healthy, PN, and MPNST patients have distinct fragment length distributions with cfDNA from MPNST being shorter in size than cfDNA from PN or healthy donors^21^, which we validated in this new cohort (**Figure 1c**). Given that plasma cfDNA comes from an admixture of cells and tissues, we hypothesized that we could more granularly leverage fragment size distributions from each NF1 peripheral nerve tumor state by applying unsupervised NMF. We therefore used NMF to deconvolve cfDNA fragment length distributions into underlying tumor versus normal fragment length signatures^31^. To accomplish this, computed cfDNA fragment length histograms from each sample were transformed into an aggregate input matrix of cfDNA fragment counts. The input matrix was considered the product of a signature matrix, representing preferred fragment lengths for each cfDNA source, and a weights matrix, representing the relative contribution of each cfDNA source to the total cfDNA admixture. Assuming two sources of cfDNA (healthy and malignant tissue), the inferred tumor-derived fragment length signature in plasma from MPNST was characteristic of previously described ctDNA with a global shift towards shorter fragment lengths and increased 10 bp periodicity (**Figure 5a**). To further ascertain whether the NMF-inferred malignant signature that we observed is derived from MPNST ctDNA, we directly compared it against ichorCNA tumor fractions. Indeed, in MPNST samples, tumor fractions in plasma measured by ichorCNA correlated strongly with NMF-inferred malignant signature weight (r = 0.6, *p* = 0.002), suggesting that NMF is separating MPNST from background cfDNA sources.

**Figure 5.**
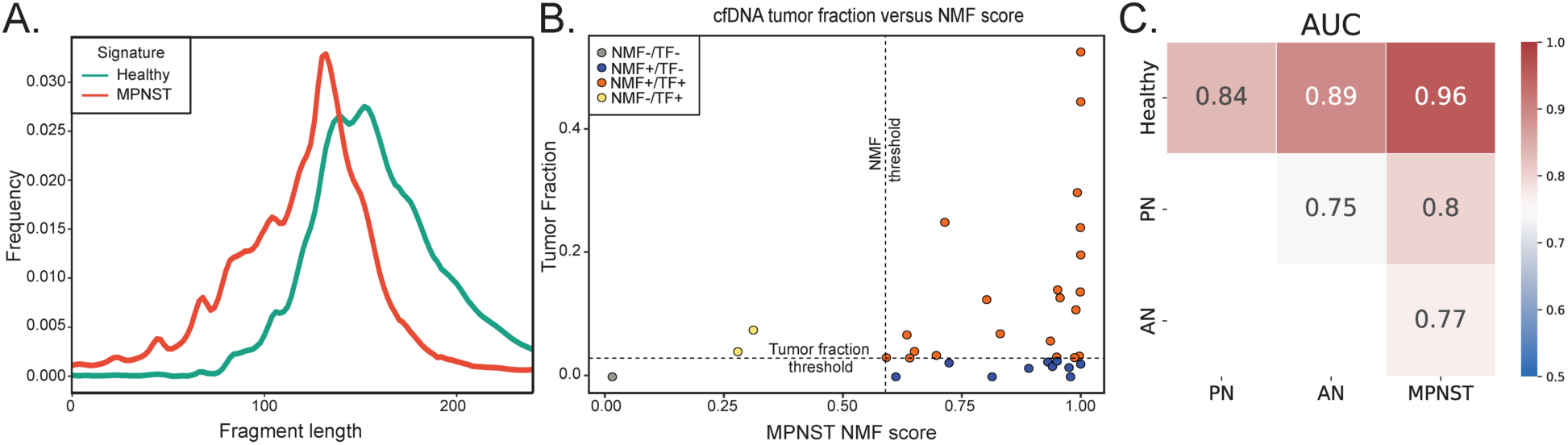
NMF decomposition of cfDNA fragment signatures distinguishes benign, pre- malignant and malignant peripheral nerve sheath tumors. (**A**) Fragment length signatures inferred from two-component NMF decomposition of healthy and MPNST cfDNA samples (**Methods**). **(B)** Correlation between CNA-derived tumor fraction and NMF MPNST score. Circle colors denote samples correctly classified by NMF and tumor fraction (red), NMF only (blue), tumor fraction only (yellow), or misclassified by both NMF and tumor fraction (gray). The thresholds for detecting MPNST by tumor fraction (healthy-MPNST 0.0305, **Methods**) and NMF (healthy-MPNST 0.5895, **Methods**) is denoted by horizontal and vertical dashed lines respectively. **(C)** ROC AUCs of logistic regression following 10 repeats of 5-fold cross-validation using one-versus-one plasma cfDNA NMF deconvolution scores as input. Healthy and disease states included within each comparison are shown along x- and y-axes, with AUCs indicated both numerically and by heat level. AN, atypical neurofibroma; AUC, area under the curve; cfDNA, cell-free DNA; CNA, copy number alteration; MPNST, malignant peripheral nerve sheath tumor; NMF, non-negative matrix factorization; PN, plexiform neurofibroma; ROC, receiver operating characteristic; TF, tumor fraction.

We next trained a logistic regression with ten repeats of fivefold cross-validation on NMF- deconvoluted weights from healthy and MPNST patient plasma and tested whether higher numbers of components could improve classification (**Supplemental Figure 4**). A logistic regression model trained on the signature weights inferred from the 20-component NMF model was able to detect tumor signal in 6 MPNST plasma samples that fell below the tumor fraction detection threshold (MPNST-healthy threshold 0.0305), with 32 of 35 (91.4%) MPNST plasma samples detectable by the NMF deconvolution approach, compared to 26 of 35 (74.3%) using ichorCNA tumor fraction (**Figure 5b)**.

Given the superior ability of fragmentomic NMF to detect MPNST from plasma, we extended this approach to the remaining disease state comparisons (**Figure 5c**). Our new approach granularly distinguished between healthy, pre-malignant and malignant disease states, including capably distinguishing between healthy and PN (AUC = 0.84), PN and AN (AUC = 0.75) and AN and MPNST (AUC = 0.77). These data suggest that NMF deconvolution applied to plasma cfDNA fragment length distributions can track disease progression across pre-malignant and malignant states in NF1, which would be clinically transformational for early cancer and pre- cancer detection for patients with hereditary cancer predisposition syndromes.

### cfDNA fragmentomics can adjudicate diagnostic challenges in patients with NF1

Having established that cfDNA fragmentomic features reliably discriminate between peripheral nerve sheath tumor disease states, we next sought to investigate whether liquid biopsy cfDNA fragmentomics could outperform conventional invasive tissue biopsy in diagnostically challenging clinical cases. A major obstacle faced by current diagnostic modalities in the early detection setting is the significant tissue heterogeneity of peripheral nerve sheath tumors^13^. Not only are benign PNs histologically heterogenous^32,33^ but, in the setting of NF1, this is further complicated by the fact that MPNST and AN often arise from directly within PN lesions^3,5^ resulting in interfacing tissues from multiple disease states.

We thus hypothesized that liquid biopsy cfDNA fragmentomics could overcome the tissue heterogeneity issue that vexes solid tumor biopsy^34,35^. To test this, we applied cfDNA fragment length NMF to Subj011, who had a diagnostic discordance between tissue biopsy and tumor resection surgical pathology (**Figure 6a**). Specifically, this patient with an FDG-avid left scapular tumor had tumor tissue biopsy consistent with AN, which guided subsequent narrow-margin surgical resection. Unfortunately, surgical pathology of the resected tissue revealed high-grade MPNST (co-occurring with AN and PN) and the narrow surgical margins, guided by the earlier AN histological diagnosis, were deemed inadequate. To adjudicate these conflicting results, we performed plasma cfDNA fragment NMF at multiple timepoints, which consistently revealed MPNST at both the biopsy and surgery. Following surgery and around the time of adjuvant radiotherapy, fragmentomic liquid biopsy MPNST signal fell below the threshold, consistent with the patient’s no evidence of disease (NED) state at that time. The patient experienced a locoregional recurrence of MPNST five months later, potentially impacted by the previous choice for narrow-margin resection. We again detected MPNST with cfDNA fragment length NMF at the same timepoint in plasma. Even looking at the NED timepoint after surgery, Subj011’s modest reduction of fragmentomic NMF score was just below the threshold.

**Figure 6.**
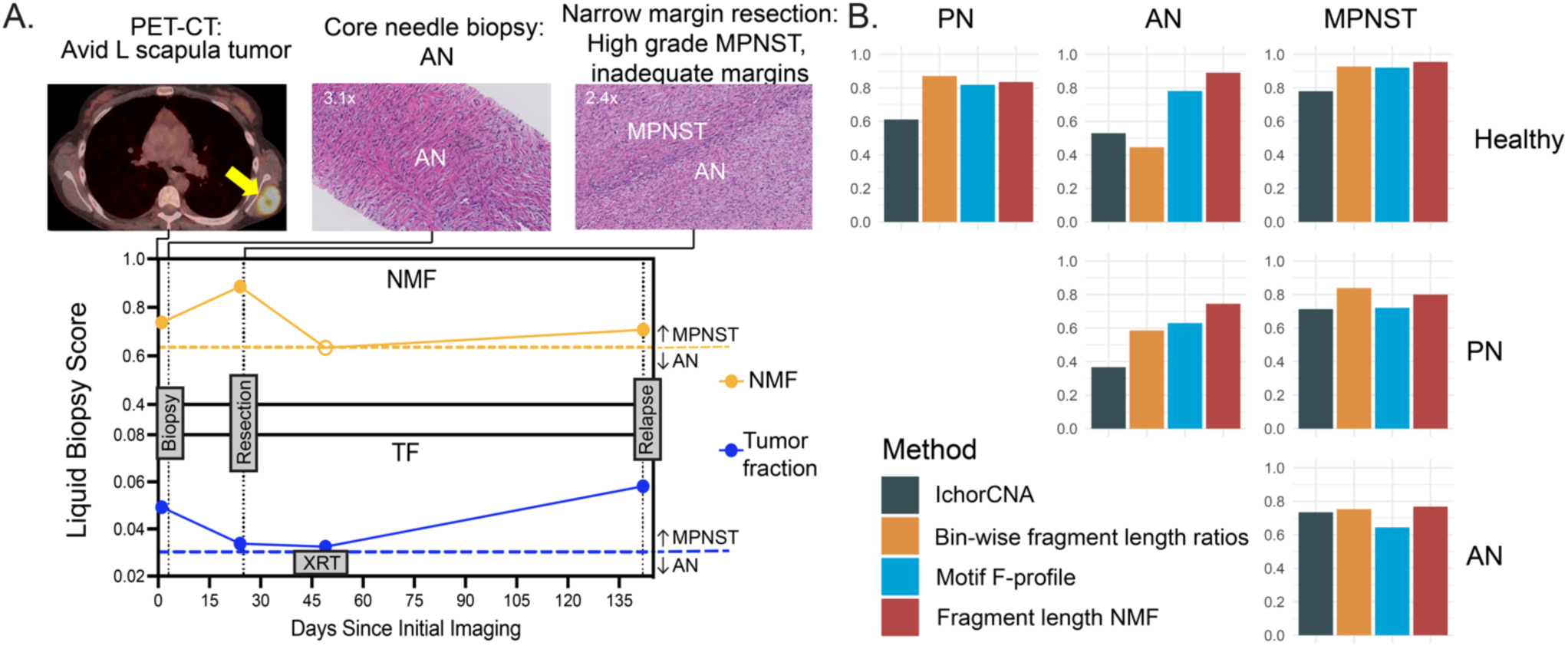
cfDNA fragment length NMF adjudicates diagnostic challenges and granularly classifies across healthy and disease states. (**A**) NMF of cfDNA fragment lengths correctly classifies MPNST initially misclassified as AN by tissue biopsy. This patient had core needle biopsy of a PET-CT avid left scapular tumor. Histopathology of the needle biopsy was consistent with a pre-malignant AN. Given the morbid location and non-malignant pathology, the patient underwent a narrow margin resection of the tumor. By NMF, both pre-biopsy and pre-resection plasma cfDNA was consistent with MPNST. Indeed, histopathology of the surgical resection tissue revealed foci of high-grade MPNST within interfacing AN and PN. Post-resection plasma cfDNA showed moderately decreased NMF scores below the MPNST threshold. Due to inadequate oncologic margins the patient underwent radiation therapy. On day 117 post-resection, the patient was found to have radiographic evidence of MPNST at the edge of the radiation field consistent with locoregional recurrence. Corresponding plasma cfDNA NMF was again consistent with MPNST. CNA derived tumor fraction from cfDNA also identified MPNST at the first and last timepoints. MPNST detection cutoffs are indicated by dashed lines with closed circles above the line indicating liquid biopsy MPNST detection. (**B**) Comparison of CNA-based and fragmentomic cell-free DNA liquid biopsy methods’ ROC AUCs across all non-malignant, pre-malignant and malignant disease states. The inset legend indicates the utilized method. Data shown are 10 repeat 5-fold cross-validated. AN, atypical neurofibroma; cfDNA, cell-free DNA; L, left; MPNST, malignant peripheral nerve sheath tumor; NMF, non-negative matrix factorization; PN, plexiform neurofibroma; TF, tumor fraction; XRT, radiation therapy.

Reassuringly, CNA-derived tumor fraction in plasma also had dynamic changes over Subj011’s clinical course (**Figure 6a**), however, liquid biopsy fragment-length NMF performed most consistently across all malignant, pre-malignant, and non-malignant disease states in this study relative to other CNA- and fragmentomic-based methodologies (**Figure 6b**).

## Discussion

To our knowledge, this study represents the most extensive collection of cfDNA data of NF1 patients to date, offering unprecedented liquid biopsy insights into the disorder. Indeed, our study heralds a paradigm shift in the early detection of MPNST, a deadly form of sarcoma that escapes modern clinical and imaging surveillance in patients with the NF1 hereditary cancer predisposition syndrome. We differentiate NF1-associated PNSTs by generating fragment-based features from cfDNA which allow us to granularly distinguish between non-malignant, pre-malignant, and frankly malignant forms of nerve sheath tumors. Our novel approach mitigates the diagnostic challenges posed by tissue heterogeneity and the concern for nerve injury with solid tumor biopsy of neurofibromas, and it offers a leap forward in personalized patient management. We anticipate that these findings will catalyze the development of non-invasive clinical assays, enabling earlier cancer detection, earlier intervention, and potentially improving the prognosis for NF1 patients at risk for MPNST. Evidenced by the recent description of the cfDNA fragmentome distinguishing patients with Li Fraumeni Syndrome (LFS) with or without cancer^36^, we anticipate the fragmentome as being a critical differentiating feature in multiple cancer predispositions. Our methodology, underscored here by the largest ever published cohort of NF1 patients profiled by cell-free DNA analysis, may therefore extend to other cancer predisposition syndromes characterized by pre-malignant tumors where early cancer detection is critical but remains elusive, often instead necessitating morbid prophylactic surgeries.

The present study advances significantly upon our previously published work in which we showed that copy number alterations in shorter cell-free DNA fragments can be used to classify MPNST versus benign PN^21^. In that study, we found that copy number alterations alone from WGS cfDNA were not robust at distinguishing MPNST from plexiform neurofibroma, but restricting our copy number analysis to shorter fragment sizes yielded a strong classifier that could distinguish these two entities from one another with high accuracy. Here we validate this finding using a completely new multi-institutional cohort of patients, again showing that size-selected cfDNA CNA analysis can distinguish MPNST from PN. This validation is a significant step forward in advancing an MPNST surveillance cfDNA assay into clinical care.

However, including AN in our validation cohort demonstrates that CNA-based methods like the one we published in 2021^21^ have limited performance in distinguishing benign PN from pre- malignant AN. This is not surprising as published tumor sequencing data report few CNAs in both AN and PN relative to MPNST^37^. Still, the distinction between PN and AN has significant clinical implications, and it will be critical to distinguish these two entities from one another if we are to implement liquid biopsy screening in the future. Clinically, asymptomatic PNs are observed with surveillance, while ANs are removed with narrow-margin resection due to their elevated risk for progression to MPNST. MPNST lesions that are localized and amenable to surgery, in contrast, are removed with wide-margin resection to reduce the risk for locoregional relapse. Therefore, a meaningful screening assay would need to granularly distinguish between all of these entities, similar in principle to colon cancer screening which requires distinction between benign polyps, high-grade polyps, and frank malignancy to have maximal screening utility^38^. Our liquid biopsy work here takes us strongly in this direction with AN vs. PN ROC AUC of 0.75, and AN vs. MPNST AUC of 0.77 using cfDNA fragment length NMF deconvolution.

Fragment length NMF methodology will need to be validated in held-out cohorts, and ultimately tested in a screening setting to fully ascertain clinical utility for detecting both pre-cancerous and cancerous lesions.

In addition to cfDNA fragment length NMF deconvolution, we also performed bin-wise fragmentomic analysis using methodology similar to Cristiano et al. and Mathios et al.^22,23^ and end motif repertoire analyses across specific motifs, motif diversity levels, and NMF deconvolution (F-profiles)^30^. All of these fragmentomic methodologies demonstrate unique but overlapping utility in distinguishing between benign, pre-cancerous, and cancerous disease states (**Figure 6b**), and there may be future utility in integrating a multi-omic fragmentomic technology for detecting pre-cancer and cancer early in NF1 patients. We elected not to do this here given that doing so would require a larger training cohort with a comparably large-sized validation cohort, a limitation given the rare nature of the hereditary cancer predisposition syndrome we studied.

In clinical vignettes, we also showcased the ability of our approach to distinguish AN from MPNST, a clinical conundrum within our standard-of-care with challenging consequences. For example, one of our patients had evidence of AN on tumor tissue biopsy, and therefore had a narrow margin resection which revealed MPNST. The patient then developed locoregional relapse shortly thereafter, perhaps related to inadequate surgical margins. Our plasma cfDNA fragmentomics approach, however, consistently detected MPNST in plasma both at the time of biopsy, the time of surgical resection, and at the time of tumor relapse months later.

Leveraging fragmentomic features in cfDNA not only enabled classification of low-mutational burden PN and AN but also improved the performance for detection of MPNST. Indeed, cfDNA fragmentomics improved the accuracy of MPNST detection relative to copy number-based tumor fraction, with NMF correctly identifying six of nine MPNST samples that were misclassified by our previously published^21^ copy number-based approach (**Figure 5b**). On the other end of the spectrum, all of the fragmentomic approaches we employed here were able to distinguish plasma from healthy volunteers from patients with benign PN, a distinction which was not possible with our previously published genome-wide copy number-based approach.

This has clinical meaning as it could enable us to detect the arc of pre-malignancy at its earliest inception point, and furthermore, could facilitate liquid biopsy approaches for tracking PN burden, which is especially important in patients with symptomatic PN on MEK inhibitors^17^ and could reduce the volume of costly and somewhat impractical whole body MRI studies.

Beyond optimizing clinical assay performance, generating a diverse fragmentomic feature set may itself provide insights into the biology of NF1. For instance, prevalence of end motifs in cfDNA reflect site-specific cutting preferences of DNases, resulting in F-profiles associated to specific DNase enzymes^30^. We observe patterns of motif enrichment in our F-profiles consistent with known DNase associations, such as enrichment of CCNN motifs in F-profile 6, a pattern associated with DNASE1L3 cutting (**Supplemental Figure 1**). In another example, we observe significantly longer fragment lengths in PN than MPNST. In other conditions, increased DNA methylation has been correlated with reduced nucleosome accessibility and subsequently impaired nuclease cutting during DNA fragmentation resulting in longer fragments^24^. MPNST, but not AN or PN, are characterized by mutations in PRC2 complex genes *EED* and *SUZ12*^37,39–41^, and loss of PRC2 function lifts transcription repression by reducing H3 lysine 27 (H3K27) methylation^42^. Therefore, longer PN fragments are consistent with intact PRC2 and increased methylation relative to MPNST. However, PN fragments in our study were also significantly longer than fragments from AN (D = 0. 0300, p < 0.001 by two-sample KS test) or healthy controls (D = 0.0241, p< 0.001 by two-sample KS test). This finding presents the intriguing possibility that global methylation is increased in PN tumors, a question not yet studied in the NF1 literature. With such fragmentomic-derived biological inferences, we look forward to a synergistic acceleration in our understanding of NF1 tumor progression as tissue informs cfDNA findings and cfDNA findings inform tissue biology.

Despite the paradigm-shifting nature of our study, it has key limitations, one of which is that it was not a prospective screening study. Ultimately, to demonstrate that our methods have clinical utility for screening and detection of MPNST and its pre-malignant precursors, a prospective, appropriately powered study will ultimately need to be performed. Another limitation is the lack of serial timepoints from each individual patient, correlated with clinical and imaging findings. We show clinical vignette examples for some cases where serial plasma was available, however, a broader set might have allowed us to further showcase potential clinical utility including minimal residual disease detection after surgical resection of MPNST. Finally, advanced imaging modalities are also being tested in this space^12,43–46^, yet we were unable to seamlessly integrate our liquid biopsy findings with imaging across the cohort due to heterogeneity of the standard-of-care diagnostic modalities employed. It will be important to test our methodology in a clinical trial setting with consistent diagnostic imaging in order to practically integrate these analytical modalities in the future.

## Methods

### Healthy controls for plasma collection

After obtaining written consent, healthy donor blood samples were obtained at a single time point from appropriately consented donors at the NIH Department of Transfusion medicine (NIH protocol NCT00001846, NIH Intramural IRB identifier 99-CC-0168) and WUSTL Clinical Translational Research Unit (WUSTL protocol NCT04354064, Washington University in St. Louis (WUSTL) School of Medicine Human Research Protection Office IRB identifiers 201903142 and 201203042). Plasma samples were collected from a total of 21 healthy volunteers (**Supplemental Table 1**). Eligibility for healthy controls included age greater than 18 years old and no known history of neoplastic or hematological disorders. Protocols are available on ClinicalTrials.gov.

### NF1 patients for plasma collection and clinical classification

This study used blood samples prospectively collected from NF1 patients with PN, AN, and MPNST tumors with the aim of distinguishing these different tumor types by plasma cfDNA analysis (**Figure 1**, **Figure 2**). Patients from the NCI and WUSTL with clinically and radiographically diagnosed PN or pathology-proven AN and MPNST were enrolled onto this multi-institutional cross-sectional study with written informed consent (NCI protocol NCT01109394, NIH Intramural IRB identifier 10C0086; NCI protocol NCT00924196, NIH Intramural IRB identifier 08C0079; WUSTL protocol NCT04354064, Washington University in St. Louis School of Medicine Human Research Protection Office IRB identifiers 201903142 and 201203042) between 2016 and 2023. Additionally, pre-treatment samples from clinical trial SARC031 were included in analysis (NCT03433183); no on-treatment samples from SARC031 were considered. NF1 status was determined clinically by consensus criteria^47^. A total of 69 PN, 17 AN, 35 untreated MPNST (minimum of 30 days washout from previous treatments), 10 MPNST on treatment and 15 resected MPNST with NED peripheral blood samples were collected (**Supplemental Tables 1-3**). AN and MPNST are designated by histological diagnoses; digital pathology from AN specimens were additionally reviewed by two external sarcoma pathologists using consensus criteria (**Supplemental Figure 2**)^48^. Atypical neurofibromatosis neoplasm with unknown biological potential (ANNUBP) were grouped with AN for analyses. Samples were considered NED by histopathological assessment and long-term event free survival; NED patients had mean follow-up of 485 days (IQR: 406 to 706 days). All patients underwent clinical management and follow-up per the standard-of-care. When available, serial blood samples were collected for vignettes (**Figure 3**, **Figure 6**) at clinically indicated patient assessments. Tissue biopsies and resections were only performed if clinically indicated and research analyses were only done if sufficient materials remained post-clinical evaluation. Tissue genomic profiling was performed by CLIA certified molecular laboratories using the TruSight Oncology 500 (TSO500; Illumina, San Diego, California) when clinically indicated (**Supplemental Figure 2**). Our study examined males and females, and similar findings are reported for both sexes. All samples were collected with informed consent for research and IRB approval in accordance with the Declaration of Helsinki. Protocols are available on ClinicalTrials.gov.

### Plasma cfDNA isolation, library construction and sequencing

To isolate cfDNA, purified plasma was thawed and cfDNA was extracted from 2 to 8 mL of plasma using the QIAamp Circulating Nucleic Acid kit (Qiagen, Hilden, Germany). Extracted DNA concentration was measured using the Qubit dsDNA High-Sensitivity Assay kit (Thermo- Fisher, Waltham, Massachusetts), and cfDNA concentration and quality were assessed using a Bioanalyzer (Agilent Technologies, Santa Clara, California). Isolated cfDNA was stored at –20°C until library preparation. cfDNA sequencing libraries were constructed from 10 to 60 ng of isolated DNA using the Kapa HyperPrep kit (Roche, Basel, Switzerland) and xGen UDI-UMI Adapters (IDT, Coralville, Iowa). All libraries were generated with 8 PCR cycles. Libraries were sequenced using 150 bp paired-end reads on a NovaSeq S4 (lllumina, San Diego, California).

### Copy number-based tumor fraction analysis from cfDNA WGS

In total, 167 cfDNA libraries were sequenced for use in this study (**Supplemental Table 1**). cfDNA whole genome sequencing (WGS) libraries were aligned to GrCh38.p13, deduplicated and filtered for standard black-listed regions^49^. Libraries were retained for analysis only if they had >80% mapped bases, <0.3% error rate, >85% unique reads, and >75% Q30 bases. One hundred sixty seven libraries met these criteria and were included in downstream analysis. An additional 110 libraries from Szymanski, et al. 2021 were included to inform the size-selected CNA-based tumor fraction held-out validation data shown in **Figure 1a**.

We also performed a technical downsampling analysis using 10 plasma samples from patients with histologically confirmed MPNST that underwent cfDNA WGS to a target depth of 25x. One sample failed quality control (<85% unique reads), therefore, 9 subjects were used for this analysis. Each subject’s 25x WGS sequencing data were aligned, deduplicated and filtered as described above. Resulting bam files were then downsampled to a normalized depth of 25x using sub-sample seed values 0-4 in samtools v1.17^50^, resulting in 5 replicates per sample.

Downsampling was repeated using seed values 0-4 to target coverages of 15x, 10x, 6x, 3x, 1x, 0.6x, 0.3x and 0.1x. Size-selected CNA-based tumor fractions were then calculated as described in Szymanski et al.^21^ at each down sampled depth and compared per-subject to the matched size-selected tumor fraction at 25x for (1) absolute deviation of mean tumor fraction from the baseline mean level and (2) statistical difference between mean tumor fraction at downsampled depth versus at 25x as assessed by Welch’s T-test (**Supplemental Figure 3**).

Size-selected tumor fraction analysis by CNA was performed as previously described^21^. Briefly, WGS reads were re-aligned to hg19 and enriched for ctDNA fragments by in silico size selection of fragment lengths between 90 and 150 bp. GC content and mappability bias correction, depth-based local copy number estimates, and copy number-based estimation of tumor fraction were then performed using the ichorCNA tool (Broad v.0.2.0)^51^.

### Bin-wise fragmentomic analysis from cfDNA WGS

To assess differences in short (100-150bp) to long (151-220 bp) cfDNA fragment length ratios across the genome, the GRCh38 reference genome was first divided into non-overlapping 5 Mb bins, excluding bins with average GC content <0.3 or average mappability <0.9. Similar to previously described methods^22,52,53^, fragment-level GC correction was performed by normalizing to a target GC distribution, by assigning fragments to 1 of 100 discrete GC strata between 0 and 1 representing each fragment’s GC content. The target distribution was set as the median GC content of the 21 healthy donor plasma samples. Profiles of GC-adjusted short-to-long cfDNA ratios were also normalized across samples to account for differences in library size. Dimensionality of the short-to-long ratio features was reduced by performing a principal component analysis within each training set and keeping only the components necessary to explain 90% of the variance between OVO disease states. Z-scores for non-acrocentric autosomal chromosomal arms were also computed from GC-adjusted residuals for short and long fragments by performing locally weighted scatterplot smoothing (LOESS regression), and center-scaling these counts by the mean and standard deviation from corresponding chromosomal arms in healthy controls^22,23,53^. Principal components accounting for 90% of variance and arm-level z-scores were then input into logistic regressions, along with bin-wise short-to-long cfDNA ratio features, for final model selection. Relative importance of individual genomic-bins and chromosomal arms in differentiating AN from MPNST were graphically represented by heatmap of principal component coefficients and eigenvalues (**Figure 3a**).

Short-to-long cfDNA fragment ratios were visualized by cohort (healthy, PN, AN, MPNST, MPNST on treatment) using per-sample normalized and GC-adjusted short-to-long cfDNA ratios mapped to bins’ genomic coordinates (**Figure 3b**).

### cfDNA end motif analysis

For cfDNA sequencing read pairs with perfect alignment with the GRCh38 reference genome, end motif analysis was performed on the four terminal nucleotides of the 5’ (forward) end of each read. Motif diversity score (MDS), which is based on Shannon entropy, was then calculated to measure end motif diversity on a scale from 0 to 1, as described by Jiang, et al. 2020^54^. Four-mer end motif frequencies were also deconvolved through non-negative matrix factorization into “founder” end motif profiles (F-profiles) using the method described by Zhou, et al. 2023^30^. Briefly, a matrix of cfDNA samples and end motif frequencies was subjected to NMF analysis using the Python library sklearn (v.1.3.2) decomposition module. The percentage contribution of each F-profile in each sample was determined by non-negative least squares regression utilizing the scipy.optimize.nnls (v.1.11.3) function in Python.

### NMF deconvolution of fragment length profiles

Fragment lengths, extracted from bam files using Picard (v4.0.1.2), and filtered to remove lengths outside the 30-700 range, were structured into a counts matrix with samples as rows and fragment lengths as columns. This counts matrix was then normalized to create a frequency matrix by normalizing rows such that they summed to one. We then used the Python sklearn library (v.1.3.2) decomposition module incorporating up to 20 components, with random initialization, multiplicative updates, and Kullback-Leibler beta divergence. The resultant component weights were used to train logistic regression models as described in the following section.This method was first tested on a two-component model with the assumption that the overall cfDNA profile is an admixture of underlying tumor and non-tumor sources. One of the components was observed to have a left-shifted peak and increased 10 bp periodicity which have been previously described as distinguishing features of tumor-derived cfDNA, and was therefore assumed to be representative of the tumor source (**Figure 5a**). This assumption was tested by correlating both the percent contribution of the tumor source component as well as the logistic regression classifier score (**Figure 5b**) with the tumor fraction obtained from ichorCNA. The percent contribution of the tumor source component was computed as the weight of the tumor source component divided by the sum of all component weights. The optimal number of components for classification, based on the area under the ROC curve of the respective logistic regression classifiers, was selected for each pairwise comparison between disease states and ranged from between 18 and 20 (components: healthy-PN: 19, healthy-AN: 19, healthy- MPNST:20, PN-AN:19, PN-MPNST:18, AN-MPNST:20; **Supplemental Figure 4**).

### Logistic regression model development and implementation

Logistic regression models from the Python sklearn library (v1.3.2) were used to analyze each set of cfDNA fragmentomic features with ten repeats of 5-fold cross-validation. The sklearn.model_select.RepeatedStratifiedKFold class was used to preserve class ratios across training folds. All models were separately applied to each cfDNA fragmentomic feature type in pair-wise comparisons between non-malignant, pre-malignant and malignant disease states. Inputs to the logistic regression models were (a) principal components of fragment ratio bins responsible for 90% of overall variance and arm-level z-scores for bin-wise fragmentomics, (b) percent contribution of F-profile 4, and (c) weights of the components derived from fragment length NMF deconvolution. Hyperparameters were optimized for regularization penalty (lambda) via grid search using sklearn.model_selection.GridSearchCV. Model coefficients from every iteration of cross-validation were saved to verify model consistency and stability. Predicted sample scores were then computed for each pair-wise comparison across non-malignant, pre- malignant and malignant disease states for each fragment feature type using the median score from the test-fold of each cross-validation repeat. ROC analysis was then used to quantify each model’s discriminative power by AUC, with the optimal threshold selected by Youden’s index.

Training and cross-validation sets included healthy (n = 21), MPNST pre-treatment (n = 35), AN (n =17) and PN (n = 69) plasma cfDNA samples. MPNST on-treatment (n = 10) and treated MPNST with NED (n = 15) cfDNA samples were held out from model training but were subsequently analyzed using the trained models (**Supplemental Table 3**). Compiled sample scores for bin-wise short/long fragment ratios, chromosomal arm-level z-scores, 4mer end motif diversity scores, and fragment length NMF scores were represented by heatmap (**Figure 2b**) using ComplexHeatmap (v.2.14.0) with supervised clustering using diagnostic cohort (MPNST, AN, PN, healthy) and with samples in each cohort ranked from highest copy number alteration derived tumor fraction to lowest.

### Power and statistical analysis

Using CNA and fragment size, we previously developed a specific and sensitive classifier for MPNST versus PN using a cohort of only 14 MPNST patients^21^. Therefore, assuming a large effect size and using Cohen’s d = 0.5 and 0.8 with an α = 0.05 and power = 0.90, we project that the sample size needed to detect differences between disease states would be n = 10 samples per group (groups= healthy controls, PN, AN, MPNST). Our category group sizes met or exceeded this estimate for all comparisons (**Supplemental Table 3**). When testing associations between plasma tumor fraction or fragmentomic features and clinical status **(Figures 1-6)**, the distributions of feature scores for each clinical status were compared by Kruskal–Wallis H test with pairwise comparisons by Dunn test. Statistical analyses were performed using *R* v.4.2.2 or Prism 10 (GraphPad Software).

## Supplemental Tables

Supplemental Table 1. Participant characteristics

Supplemental Table 2. MPNST patient characteristics

Supplemental Table 3. Details of all sequencing libraries in this study

## Supplemental Figures

Supplemental Figure 1. 4-mer end motifs across F-profiles

Supplemental Figure 2. Attributes of atypical neurofibroma samples

Supplemental Figure 3. Diminishing improvements in accuracy with deeper sequencing

Supplemental Figure 4. Number of NMF components vs AUC

## Data Availability

All data produced in the present study are available upon reasonable request to the authors.

## Acknowledgements

We are grateful to the patients and families involved in this study, to the clinical research team for collection of samples and clinical data, to the Washington University Neurofibromatosis (NF) Center, and to the NCI Center for Cancer Research Intramural Research Program. We would also like to thank the Sarcoma Alliance for Research through Collaboration (SARC) for supporting this project. Thank you to Jessica Linford and Shirin Shahsavari for providing critical feedback on the manuscript. This study utilized the computational resources of the McDonnell Genome Institute at Washington University, and the High-Performance Computing Biowulf cluster at the National Institutes of Health. Images from BioRender were used to create Figure 2.

## References

1. Mautner VF, Asuagbor FA, Dombi E, et al: Assessment of benign tumor burden by whole-body MRI in patients with neurofibromatosis 1. Neuro Oncol 10:593–8, 2008

2. Mautner VF, Hartmann M, Kluwe L, et al: MRI growth patterns of plexiform neurofibromas in patients with neurofibromatosis type 1. Neuroradiology 48:160–5, 2006

3. Beert E, Brems H, Daniels B, et al: Atypical neurofibromas in neurofibromatosis type 1 are premalignant tumors. Genes Chromosomes Cancer 50:1021–32, 2011

4. Higham CS, Dombi E, Rogiers A, et al: The characteristics of 76 atypical neurofibromas as precursors to neurofibromatosis 1 associated malignant peripheral nerve sheath tumors. Neuro Oncol 20:818–825, 2018

5. Miyamoto K, Kobayashi H, Zhang L, et al: Atypical Neurofibromatous Neoplasm with Uncertain Biologic Potential in the Posterior Mediastinum of a Young Patient with Neurofibromatosis Type 1: A Case Report. Case Rep Oncol 15:988–994, 2022

6. Evans DG, Baser ME, McGaughran J, et al: Malignant peripheral nerve sheath tumours in neurofibromatosis 1. J Med Genet 39:311–4, 2002

7. Kim A, Stewart DR, Reilly KM, et al: Malignant Peripheral Nerve Sheath Tumors State of the Science: Leveraging Clinical and Biological Insights into Effective Therapies. Sarcoma 2017:7429697, 2017

8. Uusitalo E, Rantanen M, Kallionpaa RA, et al: Distinctive Cancer Associations in Patients With Neurofibromatosis Type 1. J Clin Oncol 34:1978–86, 2016

9. Prudner BC, Ball T, Rathore R, Hirbe AC: Diagnosis and management of malignant peripheral nerve sheath tumors: Current practice and future perspectives. Neurooncol Adv 2:i40–i49, 2020

10. Gachiani J, Kim D, Nelson A, Kline D: Surgical management of malignant peripheral nerve sheath tumors. Neurosurg Focus 22:E13, 2007

11. Miller DT, Freedenberg D, Schorry E, et al: Health Supervision for Children With Neurofibromatosis Type 1. Pediatrics 143, 2019

12. Ahlawat S, Blakeley JO, Rodriguez FJ, Fayad LM: Imaging biomarkers for malignant peripheral nerve sheath tumors in neurofibromatosis type 1. Neurology 93:e1076–e1084, 2019

13. Graham DS, Russell TA, Eckardt MA, et al: Oncologic Accuracy of Image-guided Percutaneous Core-Needle Biopsy of Peripheral Nerve Sheath Tumors at a High-volume Sarcoma Center. Am J Clin Oncol 42:739–743, 2019

14. Ikuta K, Nishida Y, Sakai T, et al: Surgical Treatment and Complications of Deep-Seated Nodular Plexiform Neurofibromas Associated with Neurofibromatosis Type 1. J Clin Med 11, 2022

15. Nelson CN, Dombi E, Rosenblum JS, et al: Safe marginal resection of atypical neurofibromas in neurofibromatosis type 1. J Neurosurg:1–11, 2019

16. Dombi E, Baldwin A, Marcus LJ, et al: Activity of Selumetinib in Neurofibromatosis Type 1-Related Plexiform Neurofibromas. N Engl J Med 375:2550–2560, 2016

17. Gross AM, Wolters PL, Dombi E, et al: Selumetinib in Children with Inoperable Plexiform Neurofibromas. N Engl J Med 382:1430–1442, 2020

18. Bernthal NM, Putnam A, Jones KB, et al: The effect of surgical margins on outcomes for low grade MPNSTs and atypical neurofibroma. J Surg Oncol 110:813–6, 2014

19. Reilly KM, Kim A, Blakely J, et al: Neurofibromatosis Type 1-Associated MPNST State of the Science: Outlining a Research Agenda for the Future. J Natl Cancer Inst 109, 2017

20. Hagel C, Zils U, Peiper M, et al: Histopathology and clinical outcome of NF1- associated vs. sporadic malignant peripheral nerve sheath tumors. J Neurooncol 82:187–92, 2007

21. Szymanski JJ, Sundby RT, Jones PA, et al: Cell-free DNA ultra-low-pass whole genome sequencing to distinguish malignant peripheral nerve sheath tumor (MPNST) from its benign precursor lesion: A cross-sectional study. PLOS Medicine 18:e1003734–e1003734, 2021

22. Mathios D, Johansen JS, Cristiano S, et al: Detection and characterization of lung cancer using cell-free DNA fragmentomes. Nat Commun 12:5060, 2021

23. Cristiano S, Leal A, Phallen J, et al: Genome-wide cell-free DNA fragmentation in patients with cancer. Nature 570:385-+, 2019

24. An Y, Zhao X, Zhang Z, et al: DNA methylation analysis explores the molecular basis of plasma cell-free DNA fragmentation. Nat Commun 14:287, 2023

25. Budhraja KK, McDonald BR, Stephens MD, et al: Genome-wide analysis of aberrant position and sequence of plasma DNA fragment ends in patients with cancer. Sci Transl Med 15:eabm6863, 2023

26. Jiang P, Sun K, Peng W, et al: Plasma DNA End-Motif Profiling as a Fragmentomic Marker in Cancer, Pregnancy, and Transplantation. Cancer Discov 10:664–673, 2020

27. Jiang P, Xie T, Ding SC, et al: Detection and characterization of jagged ends of double-stranded DNA in plasma. Genome Res 30:1144–1153, 2020

28. Lo YMD, Han DSC, Jiang P, Chiu RWK: Epigenetics, fragmentomics, and topology of cell-free DNA in liquid biopsies. Science 372, 2021

29. Zhou Z, Cheng SH, Ding SC, et al: Jagged Ends of Urinary Cell-Free DNA: Characterization and Feasibility Assessment in Bladder Cancer Detection. Clin Chem 67:621–630, 2021

30. Zhou Z, Ma ML, Chan RWY, et al: Fragmentation landscape of cell-free DNA revealed by deconvolutional analysis of end motifs. Proc Natl Acad Sci U S A 120:e2220982120, 2023

31. Renaud G, Nørgaard M, Lindberg J, et al: Unsupervised detection of fragment length signatures of circulating tumor DNA using non-negative matrix factorization. eLife:e71569, 2022

32. Kershner LJ, Choi K, Wu J, et al: Multiple Nf1 Schwann cell populations reprogram the plexiform neurofibroma tumor microenvironment. JCI Insight 7, 2022

33. Peltonen J, Jaakkola S, Lebwohl M, et al: Cellular differentiation and expression of matrix genes in type 1 neurofibromatosis. Lab Invest 59:760–71, 1988

34. Pereira B, Chen CT, Goyal L, et al: Cell-free DNA captures tumor heterogeneity and driver alterations in rapid autopsies with pre-treated metastatic cancer. Nature Communications 12:3199–3199, 2021

35. Sundby RT, Pan A, Shern JF: Liquid biopsies in pediatric oncology: opportunities and obstacles. Curr Opin Pediatr, 2021

36. Wong D, Luo P, Oldfield LE, et al: Early Cancer Detection in Li-Fraumeni Syndrome with Cell-Free DNA. Cancer Discov, 2023

37. Pemov A, Hansen NF, Sindiri S, et al: Low mutation burden and frequent loss of CDKN2A/B and SMARCA2, but not PRC2, define pre-malignant neurofibromatosis type 1- associated atypical neurofibromas. Neuro Oncol, 2019

38. Montminy EM, Jang A, Conner M, Karlitz JJ: Screening for Colorectal Cancer. Med Clin North Am 104:1023–1036, 2020

39. Cortes-Ciriano I, Steele CD, Piculell K, et al: Genomic Patterns of Malignant Peripheral Nerve Sheath Tumor (MPNST) Evolution Correlate with Clinical Outcome and Are Detectable in Cell-Free DNA. Cancer Discov 13:654–671, 2023

40. Zhang X, Lou HE, Gopalan V, et al: Single-cell sequencing reveals activation of core transcription factors in PRC2-deficient malignant peripheral nerve sheath tumor. Cell Rep 40:111363, 2022

41. Lee W, Teckie S, Wiesner T, et al: PRC2 is recurrently inactivated through EED or SUZ12 loss in malignant peripheral nerve sheath tumors. Nature Genetics 46:1227–1232, 2014

42. Liu X, Liu X: PRC2, Chromatin Regulation, and Human Disease: Insights From Molecular Structure and Function. Front Oncol 12:894585, 2022

43. Derlin T, Tornquist K, Munster S, et al: Comparative effectiveness of 18F-FDG PET/CT versus whole-body MRI for detection of malignant peripheral nerve sheath tumors in neurofibromatosis type 1. Clin Nucl Med 38:e19–25, 2013

44. Ho CY, Kindler JM, Persohn S, et al: Image segmentation of plexiform neurofibromas from a deep neural network using multiple b-value diffusion data. Sci Rep 10:17857, 2020

45. Mazal AT, Ashikyan O, Cheng J, et al: Diffusion-weighted imaging and diffusion tensor imaging as adjuncts toconventional MRI for the diagnosis and management of peripheral nerve sheath tumors: current perspectives and future directions. European Radiology 29:4123–4132, 2019

46. Wei CJ, Yan C, Tang Y, et al: Computed Tomography-Based Differentiation of Benign and Malignant Craniofacial Lesions in Neurofibromatosis Type I Patients: A Machine Learning Approach. Front Oncol 10:1192, 2020

47. Legius E, Messiaen L, Wolkenstein P, et al: Revised diagnostic criteria for neurofibromatosis type 1 and Legius syndrome: an international consensus recommendation. Genet Med 23:1506-1513, 2021

48. Miettinen MM, Antonescu CR, Fletcher CDM, et al: Histopathologic evaluation of atypical neurofibromatous tumors and their transformation into malignant peripheral nerve sheath tumor in patients with neurofibromatosis 1-a consensus overview. Hum Pathol 67:1–10, 2017

49. Amemiya HM, Kundaje A, Boyle AP: The ENCODE Blacklist: Identification of Problematic Regions of the Genome. Sci Rep 9:9354, 2019

50. Danecek P, Bonfield JK, Liddle J, et al: Twelve years of SAMtools and BCFtools. Gigascience 10, 2021

51. Adalsteinsson VA, Ha G, Freeman SS, et al: Scalable whole-exome sequencing of cell-free DNA reveals high concordance with metastatic tumors. Nat Commun 8:1324, 2017

52. Cristiano S, Leal A, Phallen J, et al: Genome-wide cell-free DNA fragmentation in patients with cancer. Nature 570:385–389, 2019

53. Foda ZH, Annapragada AV, Boyapati K, et al: Detecting Liver Cancer Using Cell-Free DNA Fragmentomes. Cancer Discov 13:616–631, 2023

54. Jiang P, Sun K, Peng W, et al: Plasma DNA End-Motif Profiling as a Fragmentomic Marker in Cancer, Pregnancy, and Transplantation. Cancer Discovery 10:664–673, 2020

