## Supplemental Figures for "Early detection of malignant and pre-malignant peripheral nerve tumors using cell-free DNA fragmentomics" (from another version of this preprint: version 2, 2024-02-07)

### Supplementary Figures

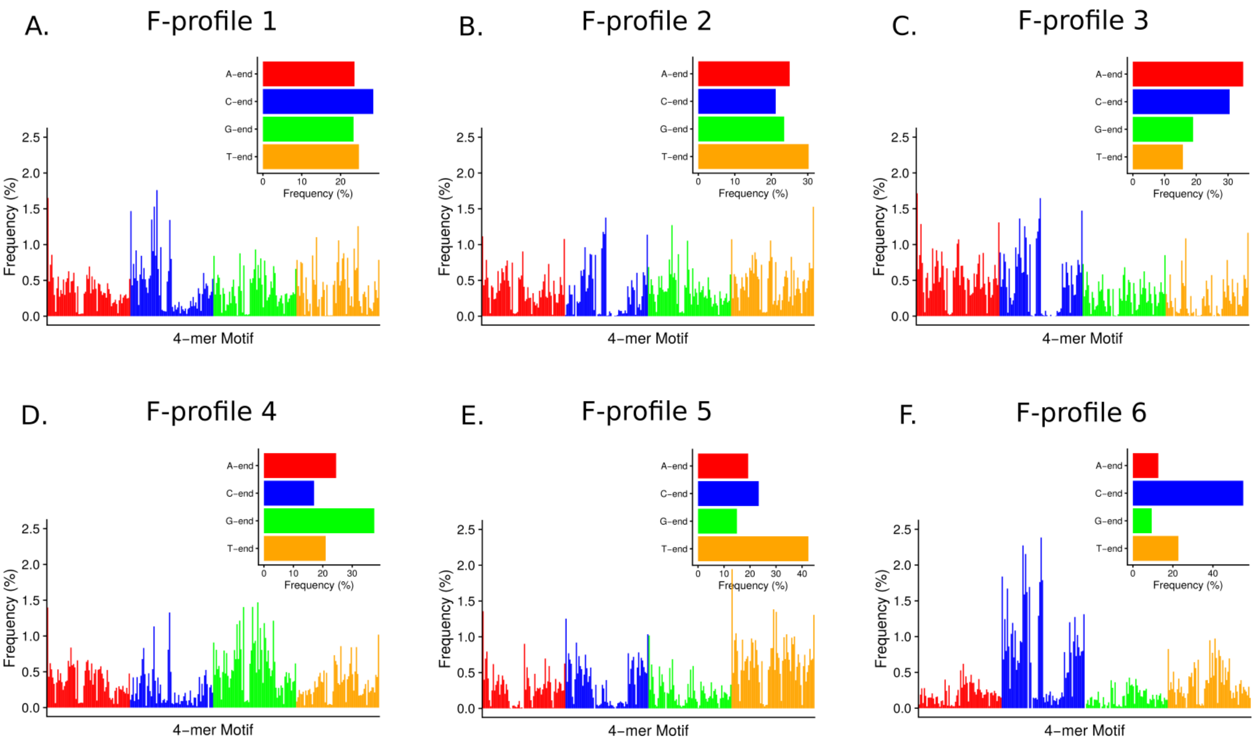


**Supplemental Figure 1. 4-mer end motifs across F-profiles**. Comprehensive view of the distribution and prevalence each possible 4-mer end motif within each of the six distinct F-profiles derived from non-negative matrix factorization (NMF) **(Methods).** The analysis highlights distinct end motif compositions and non-random distribution of end nucleotides across the F-profiles.

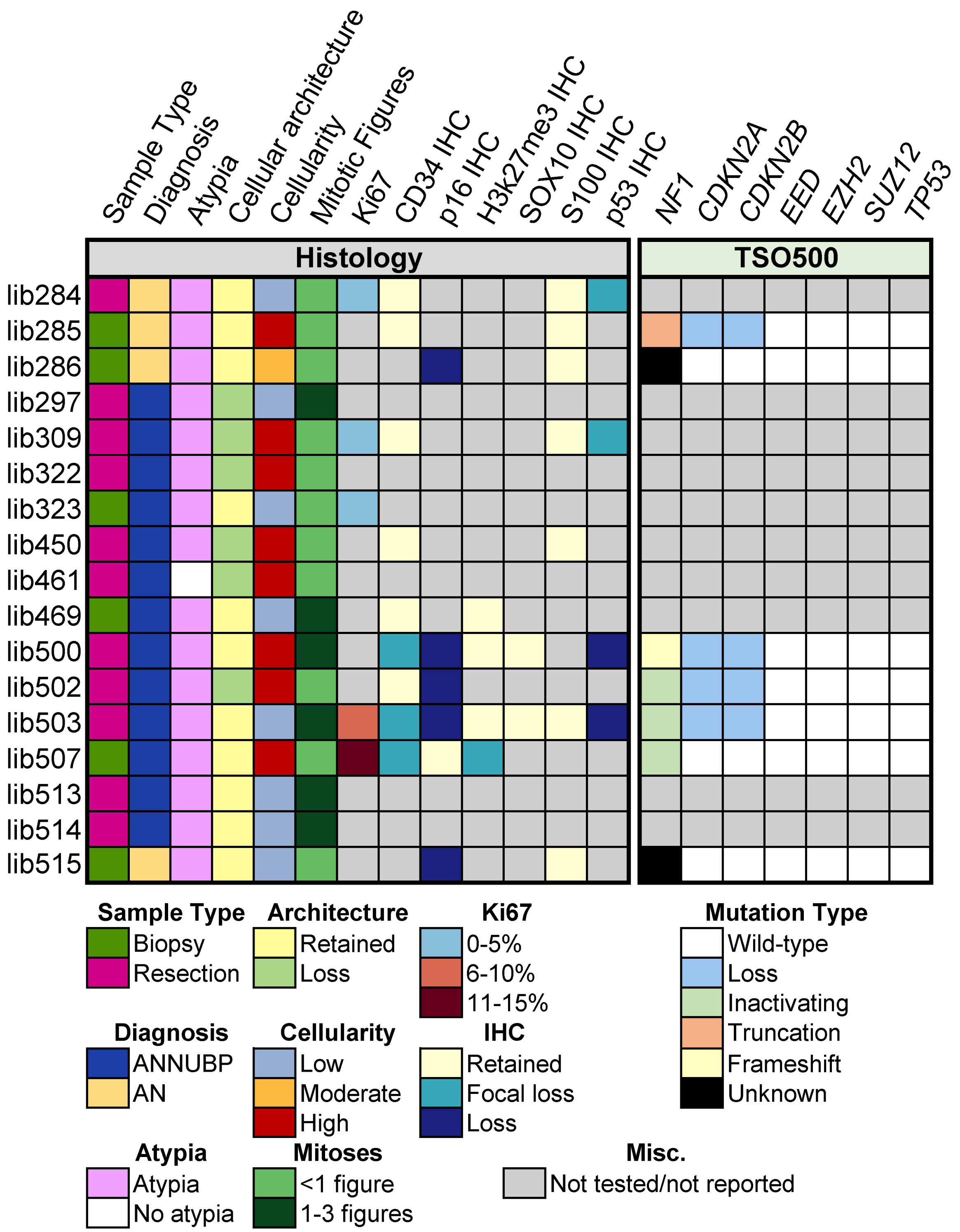


**Supplemental Figure 2. Attributes of atypical neurofibroma samples.** Histologic characteristics and, when available, TSO500 clinical sequencing results for paired tissue corresponding with AN cell-free DNA.

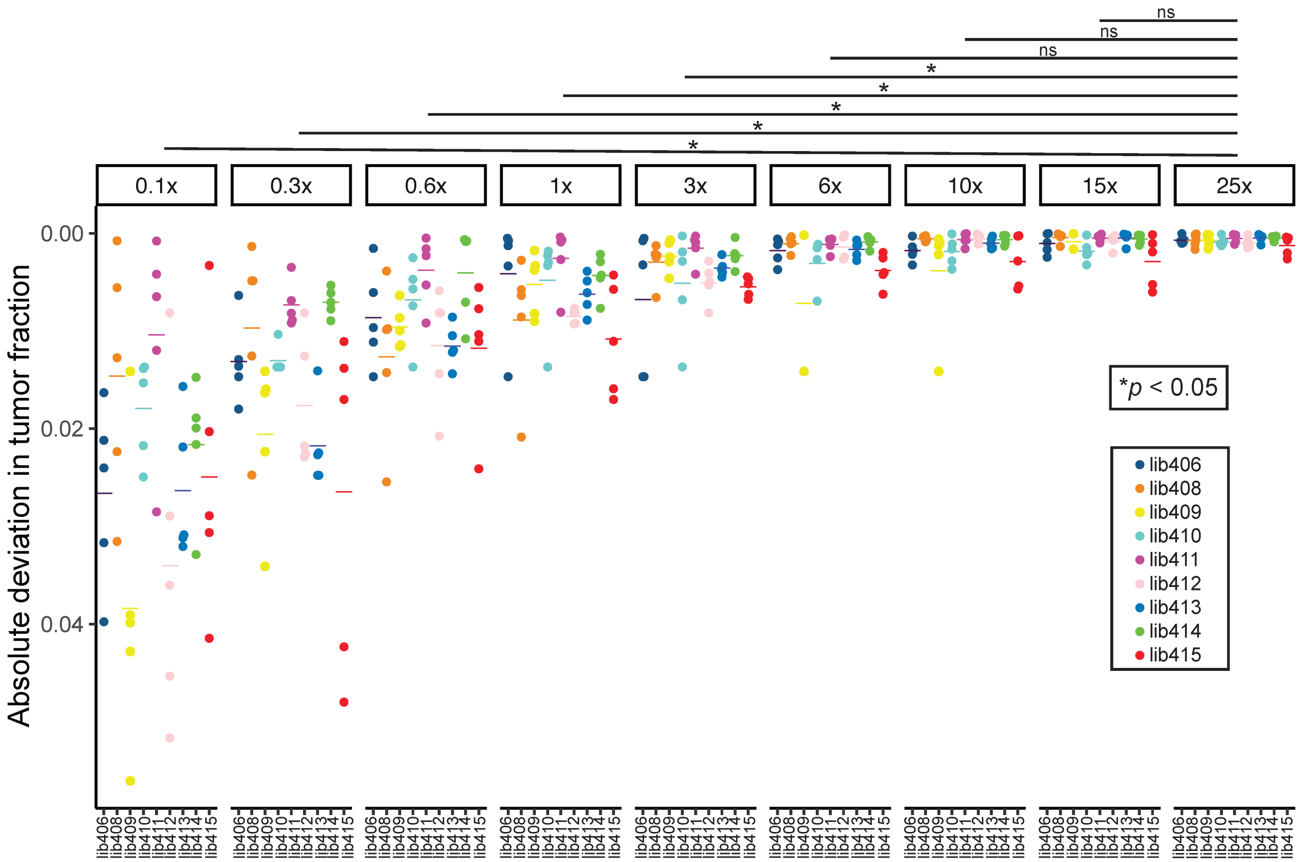


**Supplemental Figure 3. Diminishing improvements in accuracy with deeper sequencing.** MPNST samples sequenced to ~25x were downsampled in five iterations per goal coverage (**Methods**). Size selected, copy number derived tumor fraction variability was noted between iterations less than or equal to 3x with statistically significant differences in tumor fraction (*, Welch's T-test) between 25x coverage and 3x or below sequencing depths. No statistically difference (ns) was noted in tumor fraction between 25x coverage and sequencing depths greater than or equal to 6x.

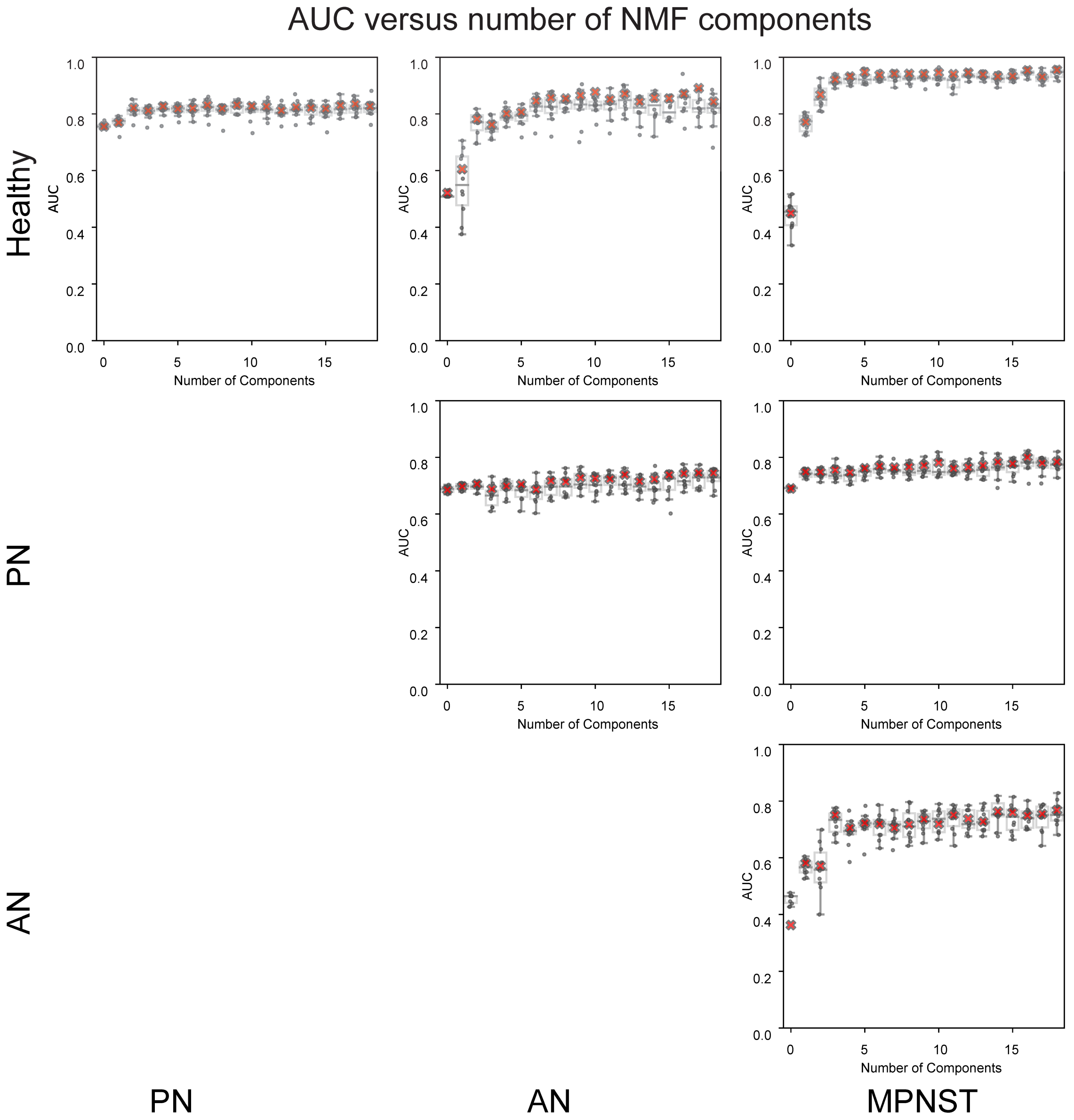


**Supplemental Figure 4. Number of NMF components versus AUC.** Performance (AUC) of NMF models in OVO comparisons is plotted against the number *n* components (**Methods**) used. The number of components approaches the AUC's asymptote in each OVO at or before 20 components. Optimal number of components for each OVO was defined as the number of components yielding the highest AUC. AUC, area under the curve; OVO, one-versus-one comparison; NMF, non-negative matrix factorization.
